# Acute Changes in Left Atrial Appendage Flow Velocity During Pulsed Field Ablation: An Intraprocedural Intracardiac Echocardiography Study

**DOI:** 10.64898/2026.10.21.26366090

**Authors:** Yu Pan, Pavel Osmančík, Tomáš Kníže, Josef Hornof, Dalibor Heřman, Sabri Hassouna, Erika Slušňáková, Štěpán Soukup, Marek Hozman

**Author notes:** Correspondence Author: Marek Hozman, MD, PhD, Department of Cardiology, Third Faculty of Medicine, Charles University and University Hospital Královské Vinohrady, Šrobárova 1150/50, 100 34 Prague 10, Czech Republic.

## Abstract

**Background:** Despite the close anatomical relationship between the left pulmonary veins (PVs) and the left atrial appendage (LAA), acute changes in LAA flow during pulsed field ablation (PFA) remain poorly characterized.

**Objectives:** To characterize acute intraprocedural changes in LAA flow and transmitral filling during PFA for atrial fibrillation (AF) using paired intracardiac echocardiography (ICE).

**Methods:** Patients with paroxysmal or persistent AF undergoing PFA were prospectively enrolled if they were in sinus rhythm at the beginning of the procedure. ICE was performed immediately before and after PFA to assess LAA emptying velocity (LAAEv), LAA filling velocity (LAAFv), transmitral E- and A-wave velocities, and E-wave deceleration time (EDT). Paired pre-to-post PFA changes were assessed, followed by a sensitivity analysis using analysis of covariance (ANCOVA) and exploratory subgroup analyses.

**Results:** Among 98 patients, LAAEv increased from 35.45 to 40.00 cm/s immediately after PFA (estimated paired increase, 4.95 cm/s; 95% CI, 2.35–7.80; P<0.001), while LAAFv increased by 4.36 cm/s (95% CI, 1.81–6.91; P=0.001). E-wave velocity increased by 8.69 cm/s and EDT decreased by 31.73 ms (both P<0.001), whereas A-wave velocity did not change significantly (P=0.08). In the ANCOVA model, changes in heart rate, systolic blood pressure, and vasopressor use were not significantly associated with changes in LAAEv. A marked acute reduction in LAAEv (≥20%) occurred in 12 patients (12.2%). No significant subgroup interactions were observed.

**Conclusions:** LAA flow velocities increased acutely after PFA, accompanied by changes in transmitral filling, although a minority of patients showed a marked acute reduction in LAA emptying.

## Introduction

Pulsed field ablation (PFA) is a nonthermal ablation technology for patients with atrial fibrillation (AF), with demonstrated procedural efficiency^1^, clinical efficacy, and safety^2, 3^. Nevertheless, several PFA-specific adverse phenomena have been reported, including coronary vasospasm^4^, intravascular haemolysis^5^, and cerebral ischaemic lesions^6–8^.

The mechanisms underlying cerebral ischaemic lesions after PFA are likely multifactorial^8, 9^. The left atrial appendage (LAA) represents an important site of blood stasis and thrombus formation in patients with AF, and reduced LAA flow velocity is associated with impaired appendage washout and thromboembolic risk^10, 11^. Given the close anatomical relationship between the LAA and the left-sided pulmonary veins^12^, PFA delivered near the left pulmonary vein ostia may acutely influence LAA mechanical function and Doppler flow velocities. Importantly, LAA flow velocities are influenced not only by appendage mechanical activity but also by atrial loading conditions and atrioventricular filling dynamics. Therefore, acute changes in LAA flow velocity may reflect a combination of local mechanical and haemodynamic effects. However, the immediate intraprocedural changes in LAA Doppler flow velocities following PFA remain poorly characterized^13^.

Transoesophageal echocardiography (TEE) is the established modality for assessing LAA function; however, its use for repeated intraprocedural measurements may be limited by the need for dedicated imaging expertise and probe manipulation^14^. In contrast, intracardiac echocardiography (ICE) is routinely used during AF ablation and allows direct imaging of the LAA during the procedure^15^. Transseptal positioning of the ICE catheter within the left atrium (LA) provides close visualization of the LAA and has been used for procedural guidance of LAA interventions^16, 17^. Visualization from the LA may be preferable for assessing LAA functional parameters, because it provides a clearer view of the ostial portion of the appendage^15, 17^.

Accordingly, the aim of the present study was to characterize immediate changes in LAA emptying velocity (LAAEv) and LAA filling velocity (LAAFv) before and after PFA in patients who were in sinus rhythm at the beginning of the procedure, thereby minimizing the potential confounding effect of rhythm conversion. ICE was used for paired intraprocedural assessment of LAA flow before and immediately after PFA. Changes in transmitral flow were also evaluated to assess acute changes in left atrioventricular filling dynamics.

## Methods

### Study Design and Population

This prospective, single-arm, single-centre observational study enrolled adults undergoing PFA for paroxysmal or persistent AF at the Cardiology Department of University Hospital Královské Vinohrady, Prague, Czech Republic.

Patients were eligible if they were aged ≥18 years, were in sinus rhythm at the beginning of the procedure, and had symptomatic AF refractory to at least one antiarrhythmic drug. Patients presenting in AF who required cardioversion to restore sinus rhythm at the beginning of the procedure were not eligible. Exclusion criteria included long-standing persistent AF, previous left atrial ablation, severe valvular heart disease or other significant structural heart disease, previous LAA closure, and pregnancy.

Patients with inadequate ICE image quality or intraprocedural atrial arrhythmia were excluded from the paired analysis. The study flow diagram, including patient screening, exclusions, and the final study population, is presented in Figure 1.

**Figure 1.**
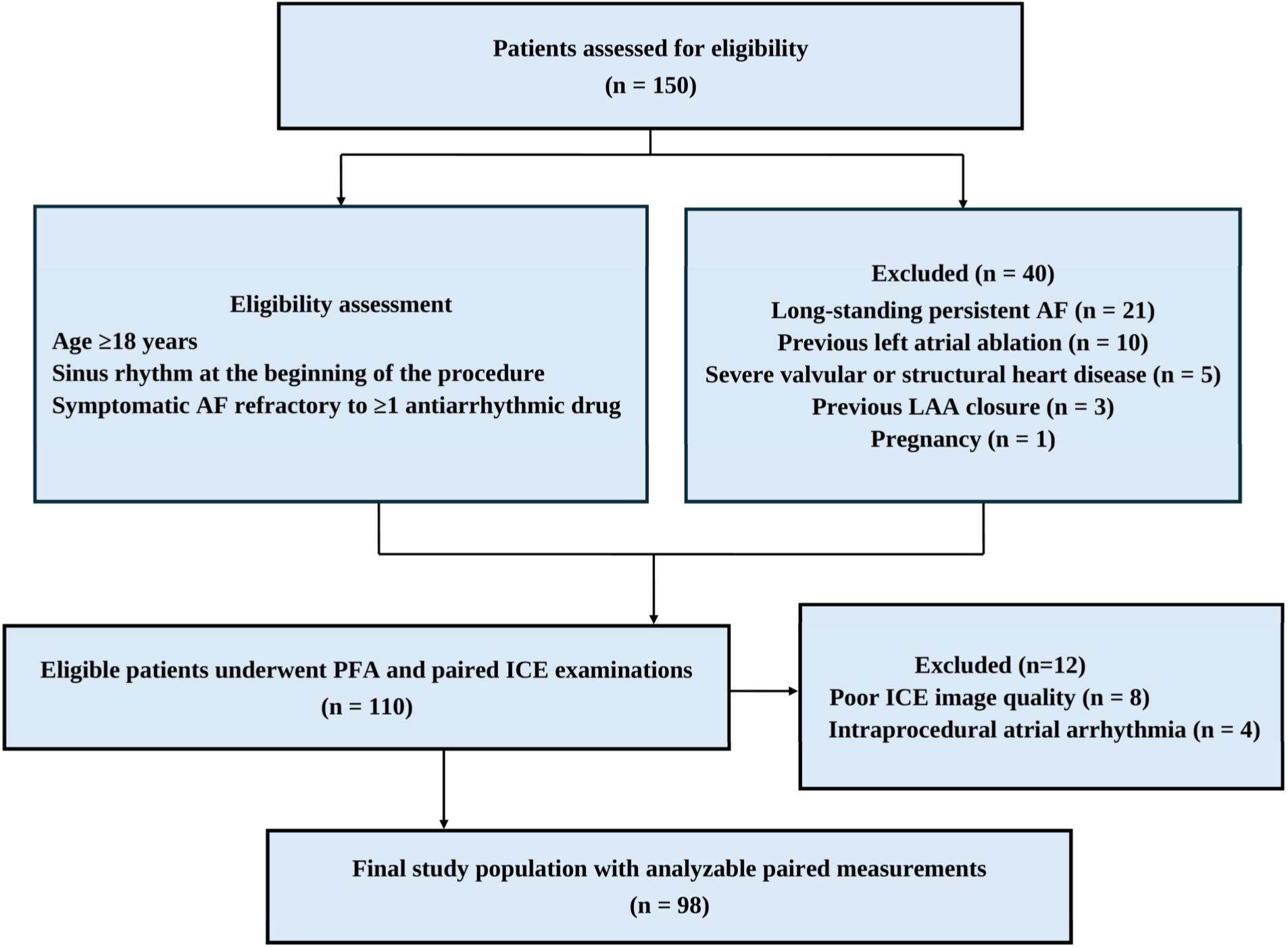
Study flow diagram. Patients assessed for eligibility, reasons for exclusion, patients undergoing PFA with paired ICE examinations, and the final study population with analysable paired measurements.

The study was conducted in accordance with the Declaration of Helsinki and was approved by the Ethics Committee for Clinical Trials of University Hospital Královské Vinohrady (approval No. EK-VP/73/00/2025). All participants provided written informed consent. The study was registered at ClinicalTrials.gov (NCT07439016).

### Pulsed Field Ablation Procedure

Procedures were performed using the FARAPULSE™ PFA system (Boston Scientific, Marlborough, MA, USA) under balanced general anaesthesia or deep sedation. All ablations were delivered using the 31-mm pentaspline PFA catheter. Pulmonary vein isolation (PVI) was performed according to the standard institutional protocol consisting of 8 pulsed field applications (4 in “basket” and 4 in “flower” configuration) per vein. Entrance block was assessed after completion of the initial PFA applications. If entrance block was not achieved, additional PFA applications were delivered until entrance block was confirmed. Exit block was also assessed, and additional PFA applications were delivered if necessary to achieve bidirectional conduction block. All procedures were guided by ICE according to institutional standards.

Additional ablation outside the pulmonary vein ostia, including the posterior wall, was performed when clinically indicated at the discretion of the treating electrophysiologist. Procedure duration, fluoroscopy time, lesion set, total number of PFA applications, and intraprocedural crystalloid volume were recorded.

### Intraprocedural Intracardiac Echocardiography

ICE was performed using a 10-Fr linear phased-array ACUSON AcuNav ICE catheter equipped with a 64-element phased-array transducer (5–10 MHz) and connected to a compatible ultrasound workstation (Siemens Healthineers, Erlangen, Germany). For pre- and post-ablation imaging, the ICE catheter was advanced into the LA through transseptal access as follows: after transseptal puncture, a guidewire was positioned in the left superior pulmonary vein, and the interatrial septum puncture site was dilated using the FARADRIVE™ sheath (Boston Scientific, Marlborough, MA, USA). The sheath was then withdrawn into the right atrium while the guidewire remained positioned within the pulmonary vein. The ICE probe was advanced into the LA via the dilated puncture site, following the guidewire under echocardiographic and fluoroscopic guidance. After the measurements were completed, the ICE probe was withdrawn to the right-sided chambers, and the transseptal sheath with the ablation catheter was placed in the LA over the guidewire. The ICE probe was reintroduced into the LA in the same manner at the end of the ablation phase. For LAA visualization, the ICE probe was positioned at the ostial aspect of the left superior pulmonary vein. The mitral valve view was obtained from the mid-LA, as described in the 2026 European Heart Rhythm Association (EHRA)/European Association of Percutaneous Cardiovascular Interventions (EAPCI) scientific statement^17^. Pulsed-wave Doppler measurements of LAAEv and LAAFv were obtained using a 3-mm sample volume positioned in the proximal portion of the LAA near its ostium. Transmitral flow velocities were recorded using a 3-mm sample volume positioned at the mitral leaflet tips.

The baseline examination was performed before the first PFA application, and repeated after completion of the planned PFA lesion set. Images were analysed offline by two investigators blinded to the measurement time point. Intraobserver and interobserver reproducibility were assessed in a randomly selected subset of 15 patients by repeating the Doppler measurements on the same stored images at both pre- and post-ablation time points. Reproducibility was quantified using intraclass correlation coefficients (ICCs). LAA Doppler measurements showed good-to-excellent reproducibility, with intraobserver ICCs of 0.954 for LAAEv and 0.885 for LAAFv and interobserver ICCs of 0.903 and 0.877, respectively (all P<0.001; Supplementary Table 1).

### LAA and Transmitral Doppler Measurements

Peak velocities of flow exiting and entering the LAA were measured using pulsed-wave Doppler and recorded as LAAEv and LAAFv, respectively (Figure 2A–B). Peak early (E wave) and late (A wave) diastolic transmitral flow velocities were measured, and the E/A ratio was calculated. E-wave deceleration time (EDT) was also measured (Figure 2C–D). For both LAA and transmitral Doppler measurements, the Doppler beam was aligned as parallel as possible to the direction of blood flow. Doppler recordings were acquired without simultaneous electrocardiographic (ECG) gating.

**Figure 2.**
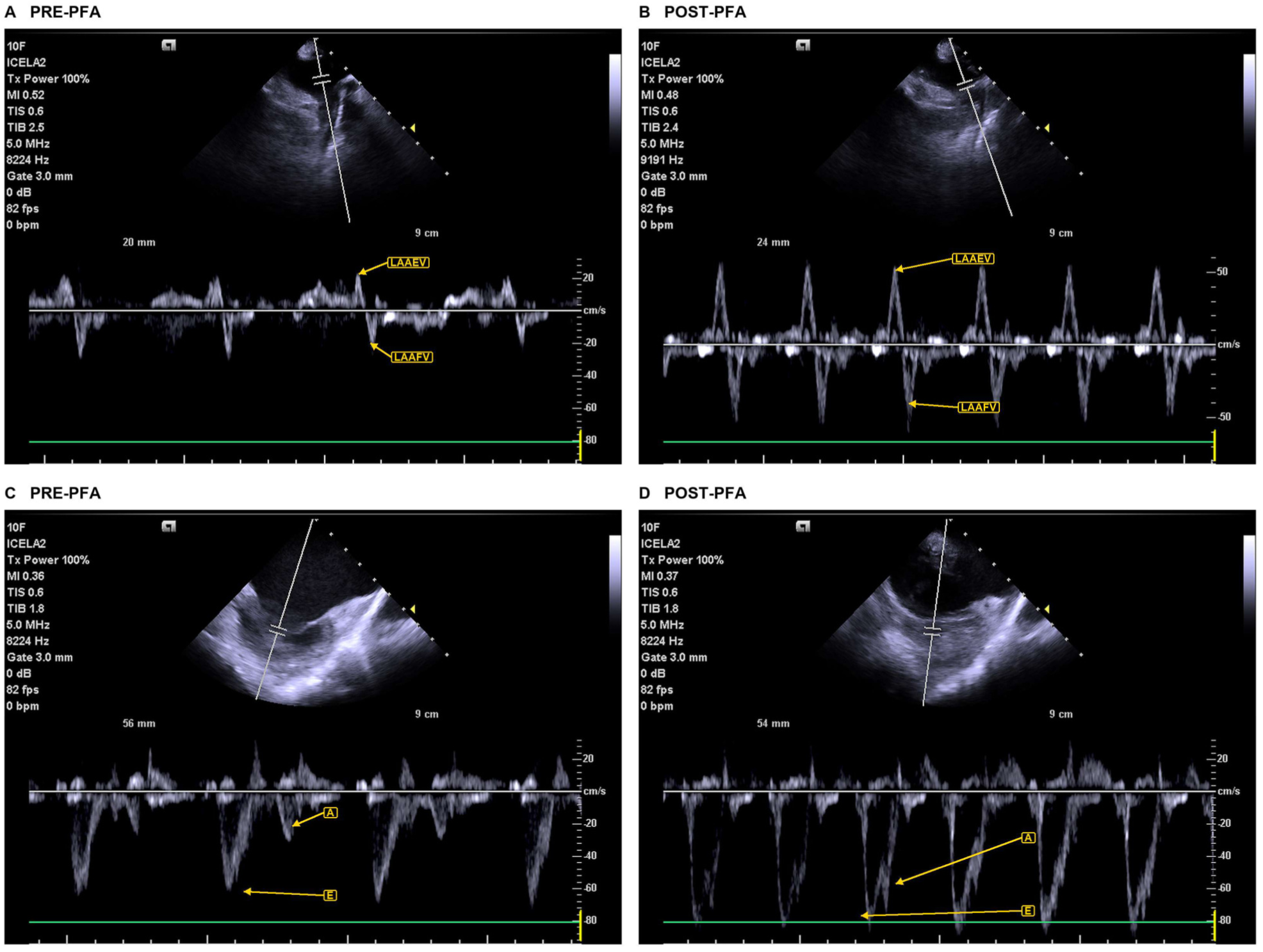
Representative intraprocedural ICE Doppler assessment before and immediately after PFA. (A, B) LAA pulsed-wave Doppler recordings showing LAA emptying and filling velocities before and after PFA. (C, D) Transmitral pulsed-wave Doppler recordings showing E- and A-wave velocities before and after PFA.

All Doppler measurements were averaged over three to five consecutive cardiac cycles.

### Anaesthesia and Intraprocedural Management

Anaesthetic management followed one of two approaches. Most patients underwent balanced general anaesthesia with a supraglottic airway or endotracheal intubation, with intravenous propofol induction, sevoflurane maintenance, and opioid analgesia. A minority (11%) of patients received total intravenous deep sedation with remimazolam and fentanyl, maintaining spontaneous breathing via a face mask, without volatile agents or propofol.

Intraprocedural vasopressors were administered as clinically indicated. Anaesthetic strategy, airway management, anaesthetic-agent use, and vasopressor administration were documented for subsequent analyses.

Intraprocedural crystalloid administration was documented and expressed as volume per kilogram of body weight. ICE images and haemodynamic measurements were acquired under the assigned anaesthetic strategy immediately before the first PFA application and after completion of the ablation procedure.

### Baseline Transthoracic Echocardiography

Comprehensive two-dimensional (2D) transthoracic echocardiography and Doppler examinations were performed one day before AF ablation using Vivid E9/E95 ultrasound systems (GE Vingmed) equipped with M5S and 4V transducers. Maximum LA volume was measured at ventricular end-systole from dedicated, nonforeshortened LA-focused apical four- and two-chamber views using the biplane method of disks and indexed to body surface area to obtain the LA volume index (LAVI)^18^. Left ventricular (LV) ejection fraction (LVEF) was calculated using the 2D biplane modified Simpson’s method^18^. Pulsed-wave tissue Doppler imaging was used to measure myocardial velocities in the basal septal and lateral segments from the apical four-chamber view.

LA longitudinal strain was assessed using two-dimensional speckle-tracking echocardiography with dedicated software (AFI LA, GE Vingmed). Images were acquired from dedicated LA-focused apical four- and two-chamber views optimized to maximize LA size and avoid foreshortening^19^. The region of interest was adjusted to follow the LA wall, excluding the pulmonary veins and LAA. The ECG R wave was used as the temporal reference for zero strain. LA reservoir, conduit, and contractile strain values were subsequently derived from the longitudinal strain curves^20^. Measurements were averaged over three to five consecutive cardiac cycles^21^.

The LA stiffness index (LASI) was calculated as the ratio of E/e′ to LA reservoir strain^22, 23^. Mean LA pressure at rest was estimated using multiparametric algorithms recommended in the 2025 American Society of Echocardiography (ASE) guidelines^24^. For examinations performed in sinus rhythm, assessment integrated mitral annular e′ velocities, E/e′, and peak tricuspid regurgitation velocity or estimated pulmonary artery systolic pressure, with E/A and supplementary parameters (pulmonary vein systolic-to-diastolic flow ratio, left atrial reservoir strain, and LAVI) incorporated as required by the algorithm.

### Study Outcomes

The primary outcome was the within-patient change in LAA emptying velocity from before to immediately after PFA. Secondary outcomes included changes in LAA filling velocity, transmitral E- and A-wave velocities, E/A ratio, and EDT.

Exploratory subgroup analyses of the primary outcome were performed according to ablation strategy (PVI alone vs. additional ablation beyond PVI), intraprocedural vasopressor use (yes vs. no), anaesthetic strategy (balanced general anaesthesia vs. deep sedation), baseline LAVI (<34 vs. ≥34 mL/m²)^21^, echocardiographically estimated left atrial pressure (LAP; normal vs. elevated)^24^, and baseline LASI (<0.42, 0.42 to <0.55, and ≥0.55). LASI cutoffs were based on published age-specific upper reference limits in healthy adults^25^.

A marked acute reduction in LAA emptying velocity was defined as a ≥20% decrease from baseline.

### Statistical Analysis

Continuous variables are presented as mean ± standard deviation (SD) or median (interquartile range [IQR]), as appropriate, and categorical variables as counts and percentages. Pre- and post-PFA measurements were compared using paired Student’s *t*-tests when the within-patient differences were normally distributed and Wilcoxon signed-rank tests otherwise. Normality of the paired differences was assessed using the Shapiro–Wilk test.

To assess whether the observed change in LAAEv could be accounted for by concomitant haemodynamic changes, an analysis of covariance (ANCOVA) was performed. The within-patient change in LAAEv (ΔLAAEv = post-PFA minus pre-PFA LAAEv) was specified as the dependent variable, with baseline LAAEv, changes in heart rate (HR; ΔHR) and systolic blood pressure (SBP; ΔSBP), and any intraprocedural vasopressor use (yes/no) included as covariates. Baseline LAAEv was mean-centred, whereas ΔHR and ΔSBP were retained on their original scales. The model intercept represented the estimated change in LAAEv at the mean baseline LAAEv, assuming no change in heart rate or systolic blood pressure and no intraprocedural vasopressor use. Heteroskedasticity-consistent type 3 (HC3) standard errors were used to calculate 95% confidence intervals (CIs) and P values.

Subgroup analyses were performed to examine whether acute changes in LAAEv differed according to procedural and anaesthesia-related factors and baseline clinical and echocardiographic characteristics. Procedural and anaesthesia-related subgroups included ablation strategy (PVI alone vs. additional ablation beyond PVI), intraprocedural vasopressor use (yes/no), and anaesthetic strategy (balanced general anaesthesia vs. deep sedation). Clinical and echocardiographic subgroups included baseline LAVI, estimated LAP, and LASI.

Differences in pre-to-post changes in LAAEv between subgroups were assessed using linear mixed-effects models with fixed effects for time, subgroup, and their interaction, and a patient-specific random intercept. Models were adjusted for age, sex, body mass index (BMI), changes in systolic blood pressure (ΔSBP) and heart rate (ΔHR), and intraprocedural crystalloid volume (mL/kg). Subgroup-specific mean changes are presented with 95% CIs, and P values for the time-by-subgroup interaction are reported as P for interaction.

In a separate exploratory analysis, a marked acute reduction in LAAEv was defined as a decrease of ≥20% from baseline. Unadjusted differences in the proportions of patients with a marked acute reduction in LAAEv across subgroup categories were compared using Fisher’s exact tests.

All tests were two-sided, with P <0.05 considered statistically significant. Analyses were performed using R version 4.6.1.

## Results

### Patient Population and Procedure Characteristics

From October 2025 to September 2026, a total of 110 patients were enrolled, and 98 of them were analysed. Twelve patients were excluded due to poor image quality or ongoing atrial arrhythmia (Figure 1). Among the 98 analysed patients, 64 (65.3%) had paroxysmal AF. All analysed patients were in sinus rhythm during the preprocedural measurement. The median age was 65 years (IQR, 57.25–72.75), and the median CHA₂DS₂-VASc score was 2.0 (1.0–3.0). Hypertension was the most common comorbidity (72.4%), followed by heart failure (21.4%) and diabetes mellitus (19.4%). Patients with persistent AF were older and had more pronounced structural and functional abnormalities on echocardiography than those with paroxysmal AF. Baseline characteristics are summarized in Table 1.

**Table 1.** Clinical, echocardiographic, and procedural characteristics of the study population. Data are presented as mean ± SD, median (IQR), or n (%), as appropriate. Percentages are based on available data for each variable. Anaesthetic strategy, airway management, and intraprocedural medication variables are presented as n/N (%). LASI was calculated as E/e′ divided by LASr. Abbreviations: AF, atrial fibrillation; BMI, body mass index; BSA, body surface area; CHA₂DS₂-VASc, congestive heart failure, hypertension, age ≥75 years, diabetes mellitus, stroke/transient ischaemic attack, vascular disease, age 65–74 years, and sex category; DM, diabetes mellitus; CAD, coronary artery disease; HF, heart failure; CKD, chronic kidney disease; TIA, transient ischaemic attack; COPD, chronic obstructive pulmonary disease; OSA, obstructive sleep apnoea; LAVI, left atrial volume index; LVEF, left ventricular ejection fraction; LASr, left atrial reservoir strain; LAEF, left atrial emptying fraction; E, early diastolic transmitral flow velocity; A, late diastolic transmitral flow velocity; EDT, E-wave deceleration time; e′, early diastolic mitral annular velocity; LASI, left atrial stiffness index; MR, mitral regurgitation; TR, tricuspid regurgitation; sPAP, systolic pulmonary artery pressure; PVI, pulmonary vein isolation; PV, pulmonary vein; PFA, pulsed field ablation; PW, posterior wall; CTI, cavotricuspid isthmus.

| Characteristic | Overall (N = 98) | Paroxysmal AF (n = 64) | Persistent AF (n = 34) |
| --- | --- | --- | --- |
| **Clinical characteristics** |  |  |  |
| Age, years | 65.00 (57.25–72.75) | 64.00 (52.75–70.00) | 71.00 (61.00–76.75) |
| BMI, kg/m<sup>2</sup> | 28.95 (26.20–32.00) | 28.55 (26.15–31.20) | 29.35 (26.22–34.50) |
| BSA, m<sup>2</sup> | 2.09 ± 0.24 | 2.09 ± 0.23 | 2.10 ± 0.26 |
| Male sex, n (%) | 62 (63.3%) | 40 (62.5%) | 22 (64.7%) |
| AF duration, months | 12.00 (7.00–36.00) | 10.00 (6.50–33.50) | 15.00 (7.75–44.75) |
| CHA<sub>2</sub>DS<sub>2</sub>-VASc score | 2.00 (1.00–3.00) | 2.00 (1.00–3.00) | 3.00 (2.00–4.00) |
| Hypertension, n (%) | 71 (72.4%) | 45 (70.3%) | 26 (76.5%) |
| DM, n (%) | 19 (19.4%) | 9 (14.1%) | 10 (29.4%) |
| CAD, n (%) | 17 (17.3%) | 8 (12.5%) | 9 (26.5%) |
| HF, n (%) | 21 (21.4%) | 6 (9.4%) | 15 (44.1%) |
| CKD, n (%) | 10 (10.2%) | 3 (4.7%) | 7 (20.6%) |
| Previous stroke/TIA, n (%) | 8 (8.2%) | 4 (6.2%) | 4 (11.8%) |
| COPD, n (%) | 8 (8.3%) | 5 (8.1%) | 3 (8.8%) |
| OSA, n (%) | 4 (4.1%) | 3 (4.7%) | 1 (2.9%) |
| **Baseline transthoracic echocardiographic characteristics** |  |  |  |
| LAVI, mL/m<sup>2</sup> | 34.75 (27.56–41.82) | 33.79 (26.08–40.00) | 40.55 (32.27–51.85) |
| LVEF, % | 60.00 (55.00–62.00) | 60.00 (58.00–63.00) | 56.50 (53.00–60.00) |
| LASr, % | 24.48 (19.25–33.75) | 29.50 (20.25–35.75) | 20.50 (17.00–25.00) |
| LAEF, % | 52.83 ± 12.27 | 55.42 ± 11.52 | 47.81 ± 12.29 |
| E, cm/s | 60.50 (51.75–75.25) | 60.00 (51.25–73.75) | 62.50 (53.25–81.00) |
| A, cm/s | 59.35 ± 22.62 | 60.74 ± 21.61 | 56.79 ± 24.53 |
| E/A ratio | 1.04 (0.80–1.47) | 1.03 (0.77–1.46) | 1.05 (0.82–1.50) |
| EDT, ms | 201.00 (170.00–244.00) | 209.87 (176.75–250.50) | 183.50 (158.00–225.75) |
| Mean e', cm/s | 7.89 ± 2.04 | 8.22 ± 2.11 | 7.27 ± 1.75 |
| E/e' | 8.00 (6.72–9.63) | 7.67 (6.67–9.29) | 8.67 (6.92–10.75) |
| LASI | 0.32 (0.21–0.48) | 0.27 (0.18–0.40) | 0.37 (0.27–0.62) |
| MR ≥ mild, n (%) | 27 (27.8%) | 16 (25.0%) | 11 (33.3%) |
| TR present, n (%) | 61 (62.2%) | 39 (60.9%) | 22 (64.7%) |
| sPAP, mmHg | 29.76 ± 8.36 | 28.11 ± 5.14 | 32.45 ± 11.73 |
| **Anaesthesia and intraprocedural management** |  |  |  |
| ***Anaesthetic strategy*** |  |  |  |
| Balanced general anaesthesia, n/N (%) | 87/98 (88.8%) | 54/64 (84.4%) | 33/34 (97.1%) |
| Deep sedation with spontaneous breathing, n/N (%) | 11/98 (11.2%) | 10/64 (15.6%) | 1/34 (2.9%) |
| ***Airway management*** |  |  |  |
| Supraglottic airway device, n/N (%) | 81/98 (82.7%) | 48/64 (75.0%) | 33/34 (97.1%) |
| Endotracheal intubation, n/N (%) | 6/98 (6.1%) | 6/64 (9.4%) | 0/34 (0.0%) |
| Face mask, n/N (%) | 11/98 (11.2%) | 10/64 (15.6%) | 1/34 (2.9%) |
| Propofol administration, n/N (%) | 87/98 (88.8%) | 54/64 (84.4%) | 33/34 (97.1%) |
| Volatile anaesthetic administration, n/N (%) | 87/98 (88.8%) | 54/64 (84.4%) | 33/34 (97.1%) |
| Remimazolam administration, n/N (%) | 11/98 (11.2%) | 10/64 (15.6%) | 1/34 (2.9%) |
| Opioid administration, n/N (%) | 98/98 (100.0%) | 64/64 (100.0%) | 34/34 (100.0%) |
| Vasopressor administration, n/N (%) | 47/98 (48.0%) | 25/64 (39.1%) | 22/34 (64.7%) |
| Inotrope administration, n/N (%) | 0/98 (0.0%) | 0/64 (0.0%) | 0/34 (0.0%) |
| Intraprocedural crystalloid volume, mL/kg | 5.62 (4.76–7.27) | 5.88 (4.74–7.49) | 5.41 (4.80–6.43) |
| **Procedural characteristics** |  |  |  |
| Additional ablation beyond PVI, n (%) | 35 (35.7%) | 6 (9.4%) | 29 (85.3%) |
| Procedure time, min | 46.50 (40.00–55.00) | 45.00 (40.00–50.00) | 52.00 (44.50–65.00) |
| PV PFA applications, n | 34.00 (32.00–38.00) | 34.00 (32.00–40.00) | 34.00 (32.00–36.00) |
| PW PFA applications, n | 0.00 (0.00–21.50) | 0.00 (0.00–0.00) | 28.00 (20.00–31.50) |
| CTI PFA applications, n | 0.00 (0.00–0.00) | 0.00 (0.00–0.00) | 0.00 (0.00–0.00) |
| Total PFA applications, n | 40.00 (34.00–55.75) | 36.00 (32.00–40.00) | 60.00 (54.25–67.00) |
| Fluoroscopy time, min | 4.54 (3.30–6.16) | 4.17 (3.10–5.29) | 5.45 (4.23–7.37) |

All procedures were performed under one of two anaesthetic strategies. 87 patients (88.8%) received balanced general anaesthesia with a supraglottic airway (n = 81, 82.7%) or endotracheal intubation (n = 6, 6.1%), whereas 11 patients (11.2%) received deep sedation with spontaneous breathing via a face mask. Intraprocedural vasopressors were used in 47 patients (48.0%), whereas no patients received inotropic agents. The median intraprocedural crystalloid volume was 5.62 mL/kg (IQR, 4.76–7.27). The median procedure duration was 46.5 min, the median number of PFA applications was 40, and the median fluoroscopy time was 4.54 min. Additional ablation beyond PVI was performed in 35 patients (35.7%) and was more frequent in patients with persistent AF. No serious complications occurred.

### Acute Changes in Left Atrial Appendage Function and Transmitral Flow

Intraprocedural changes in LAA flow, transmitral flow, blood pressure, and heart rate before and immediately after PFA are presented in Figure 3 and Table 2. Following PFA, LAAEv increased from 35.45 (IQR, 25.67–45.20) cm/s to 40.00 (IQR, 31.95–49.95) cm/s, with an estimated paired increase of 4.95 cm/s (95% CI, 2.35–7.80; P<0.001). LAAFv also increased, with a mean change of 4.36 cm/s (95% CI, 1.81–6.91; P=0.001) (Figure 3A and Table 2).

**Figure 3.**
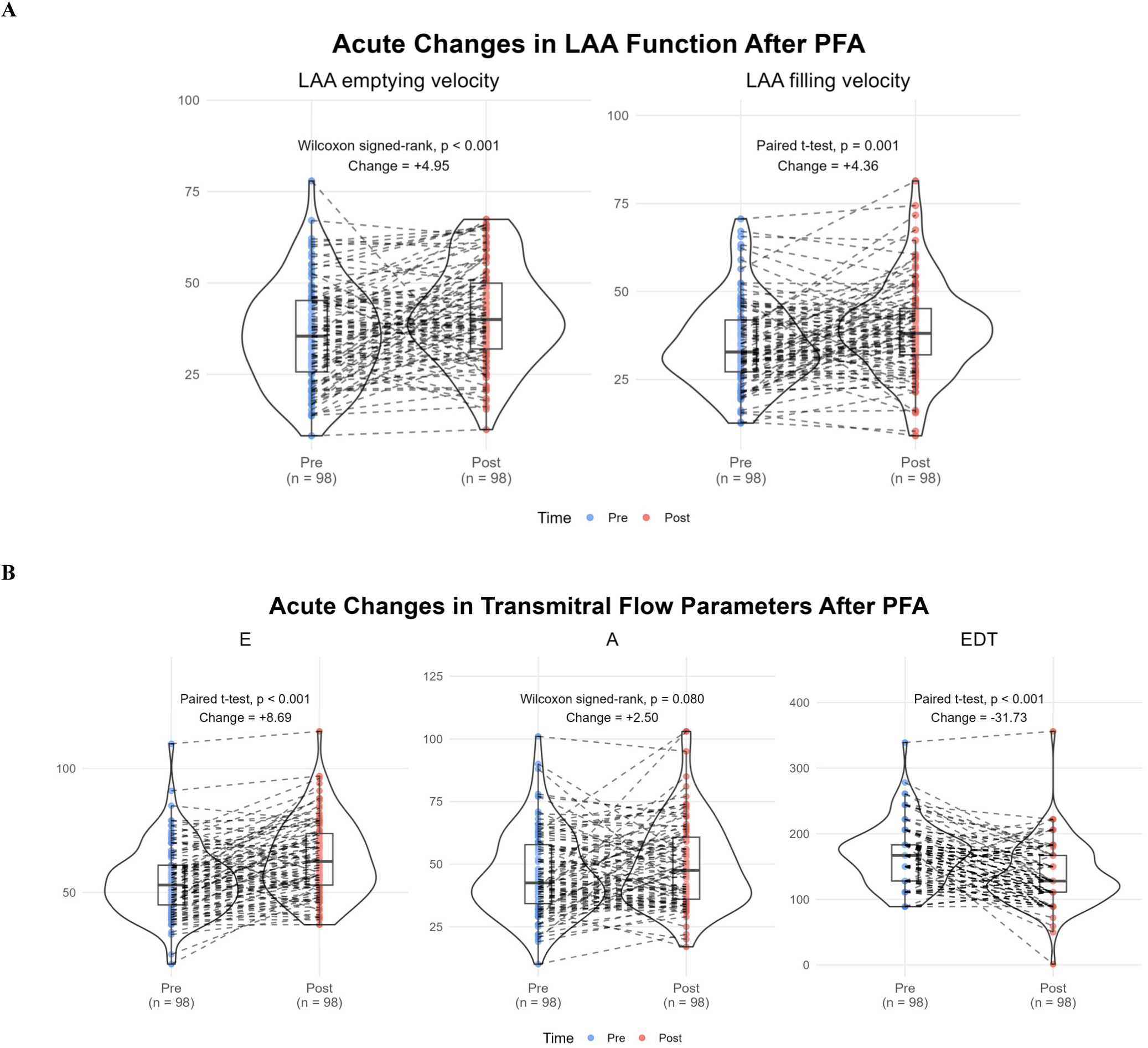
Acute changes in LAA function and transmitral Doppler parameters immediately after PFA. (A) Paired pre- and post-PFA LAA emptying and filling velocities. (B) Paired changes in transmitral E-wave velocity, A-wave velocity, and E-wave deceleration time. P values were derived from paired t tests or Wilcoxon signed-rank tests, according to the distribution of paired differences.

**Table 2.**
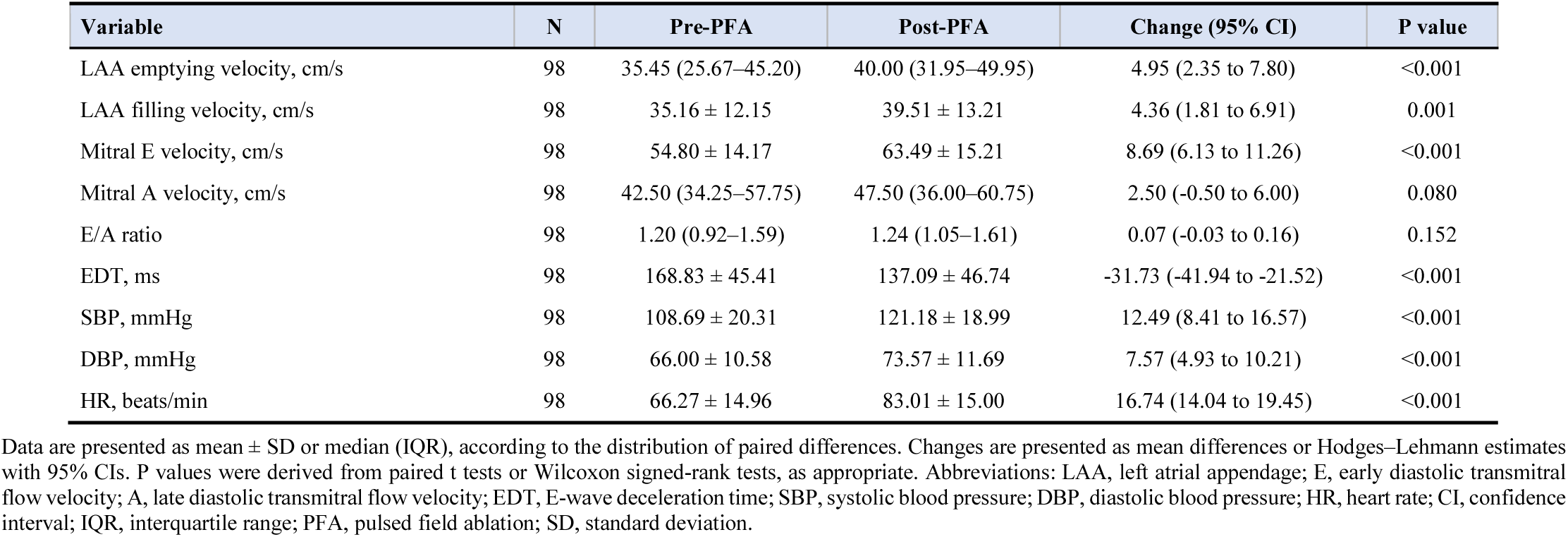
Acute changes in LAA function, transmitral flow, and haemodynamic parameters following PFA. Data are presented as mean ± SD or median (IQR), according to the distribution of paired differences. Changes are presented as mean differences or Hodges–Lehmann estimates with 95% CIs. P values were derived from paired t tests or Wilcoxon signed-rank tests, as appropriate. Abbreviations: LAA, left atrial appendage; E, early diastolic transmitral flow velocity; A, late diastolic transmitral flow velocity; EDT, E-wave deceleration time; SBP, systolic blood pressure; DBP, diastolic blood pressure; HR, heart rate; CI, confidence interval; IQR, interquartile range; PFA, pulsed field ablation; SD, standard deviation.

| Variable | N | Pre-PFA | Post-PFA | Change (95% CI) | P value |
| --- | --- | --- | --- | --- | --- |
| LAA emptying velocity, cm/s | 98 | 35.45 (25.67–45.20) | 40.00 (31.95–49.95) | 4.95 (2.35 to 7.80) | <0.001 |
| LAA filling velocity, cm/s | 98 | 35.16 ± 12.15 | 39.51 ± 13.21 | 4.36 (1.81 to 6.91) | 0.001 |
| Mitral E velocity, cm/s | 98 | 54.80 ± 14.17 | 63.49 ± 15.21 | 8.69 (6.13 to 11.26) | <0.001 |
| Mitral A velocity, cm/s | 98 | 42.50 (34.25–57.75) | 47.50 (36.00–60.75) | 2.50 (-0.50 to 6.00) | 0.080 |
| E/A ratio | 98 | 1.20 (0.92–1.59) | 1.24 (1.05–1.61) | 0.07 (-0.03 to 0.16) | 0.152 |
| EDT, ms | 98 | 168.83 ± 45.41 | 137.09 ± 46.74 | -31.73 (-41.94 to -21.52) | <0.001 |
| SBP, mmHg | 98 | 108.69 ± 20.31 | 121.18 ± 18.99 | 12.49 (8.41 to 16.57) | <0.001 |
| DBP, mmHg | 98 | 66.00 ± 10.58 | 73.57 ± 11.69 | 7.57 (4.93 to 10.21) | <0.001 |
| HR, beats/min | 98 | 66.27 ± 14.96 | 83.01 ± 15.00 | 16.74 (14.04 to 19.45) | <0.001 |

**Table 3.**
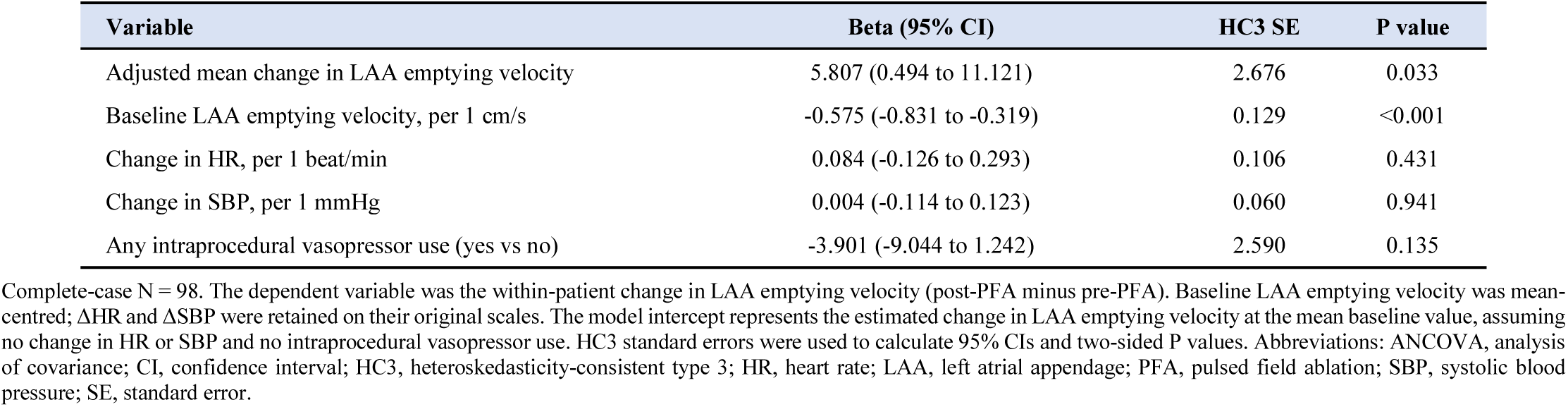
Haemodynamic ANCOVA of the acute change in LAA emptying velocity. Complete-case N = 98. The dependent variable was the within-patient change in LAA emptying velocity (post-PFA minus pre-PFA). Baseline LAA emptying velocity was mean-centred; ΔHR and ΔSBP were retained on their original scales. The model intercept represents the estimated change in LAA emptying velocity at the mean baseline value, assuming no change in HR or SBP and no intraprocedural vasopressor use. HC3 standard errors were used to calculate 95% CIs and two-sided P values. Abbreviations: ANCOVA, analysis of covariance; CI, confidence interval; HC3, heteroskedasticity-consistent type 3; HR, heart rate; LAA, left atrial appendage; PFA, pulsed field ablation; SBP, systolic blood pressure; SE, standard error.

| Variable | Beta (95% CI) | HC3 SE | P value |
| --- | --- | --- | --- |
| Adjusted mean change in LAA emptying velocity | 5.807 (0.494 to 11.121) | 2.676 | 0.033 |
| Baseline LAA emptying velocity, per 1 cm/s | -0.575 (-0.831 to -0.319) | 0.129 | <0.001 |
| Change in HR, per 1 beat/min | 0.084 (-0.126 to 0.293) | 0.106 | 0.431 |
| Change in SBP, per 1 mmHg | 0.004 (-0.114 to 0.123) | 0.060 | 0.941 |
| Any intraprocedural vasopressor use (yes vs no) | -3.901 (-9.044 to 1.242) | 2.590 | 0.135 |

Concomitant changes were observed in transmitral flow. Mitral E-wave velocity increased by 8.69 cm/s (P<0.001), whereas EDT decreased by 31.73 ms (P<0.001). In contrast, mitral A-wave velocity and the E/A ratio did not change significantly (P=0.08 and P=0.15, respectively) (Figure 3B and Table 2).

### Sensitivity Analysis Accounting for Haemodynamic Changes

In the multivariable ANCOVA model adjusting for baseline LAAEv, changes in heart rate (ΔHR) and systolic blood pressure (ΔSBP), and intraprocedural vasopressor use, the estimated change in LAAEv was 5.81 cm/s (95% CI, 0.49–11.12; P = 0.033) at the mean baseline LAAEv, assuming no change in HR or SBP and no vasopressor use. Baseline LAAEv was inversely associated with ΔLAAEv (β = −0.575; 95% CI, −0.831 to −0.319; P < 0.001). Neither ΔHR (β = 0.084 per bpm; P = 0.431), ΔSBP (β = 0.004 per mmHg; P = 0.941), nor vasopressor use (β = −3.901; P = 0.135) was significantly associated with ΔLAAEv.

### Subgroup Analyses of Acute Changes in Left Atrial Appendage Emptying Velocity

Subgroup analyses of the mean change in LAAEv and the incidence of marked acute reduction are presented in Figures 4 and 5. The mean increase in LAAEv was comparable between patients undergoing PVI alone and those receiving additional ablation beyond PVI (5.5 vs. 5.1 cm/s; P for interaction = 0.671). Patients who did not receive intraprocedural vasopressors showed a numerically greater increase than those receiving vasopressors (7.4 vs. 3.2 cm/s; P for interaction = 0.377). Mean changes were 5.2 and 6.5 cm/s in patients receiving balanced general anaesthesia and intravenous deep sedation, respectively (P for interaction = 0.990).

**Figure 4.**
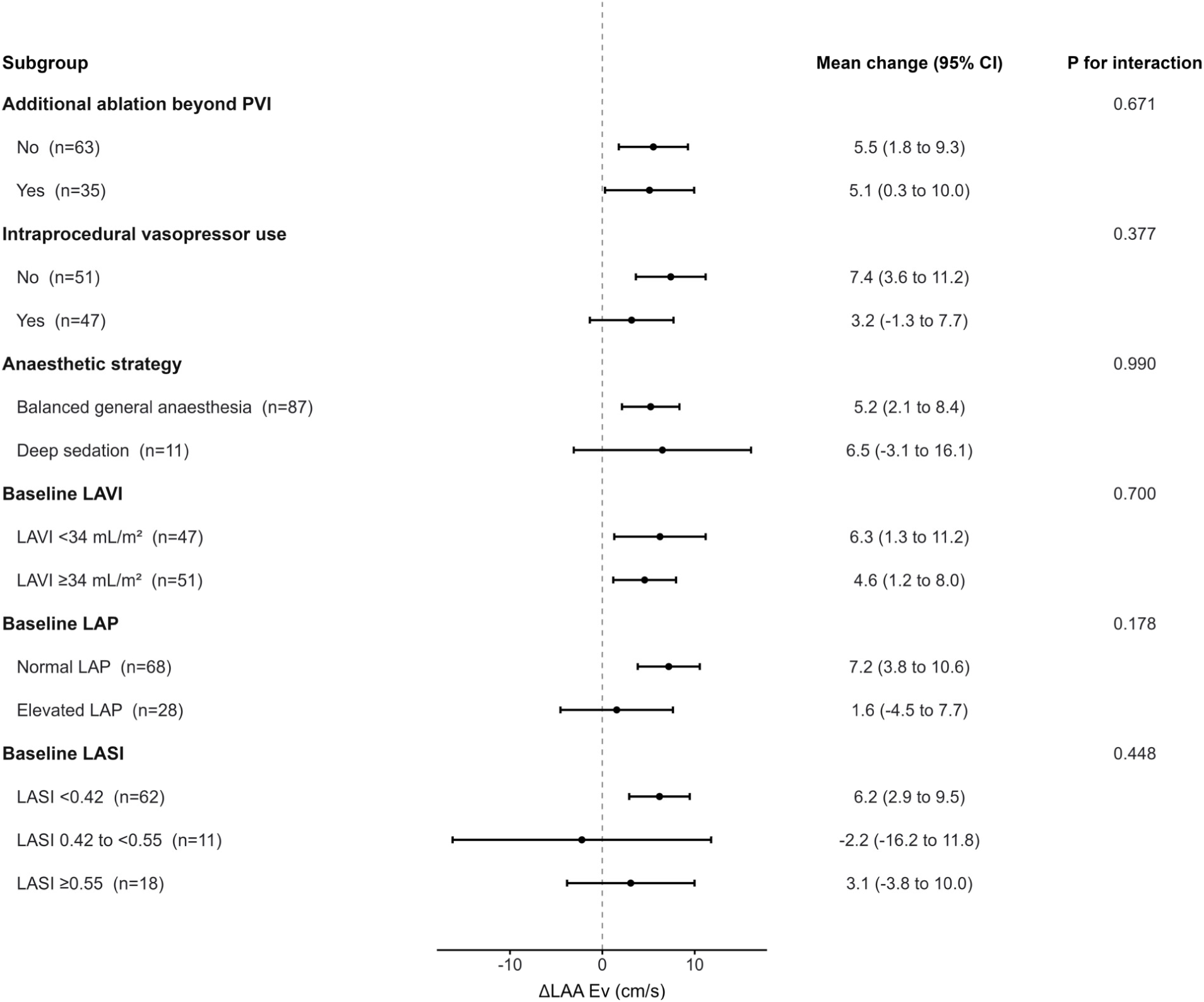
Subgroup analyses of the acute change in LAA emptying velocity after PFA. Points and horizontal lines indicate observed subgroup-specific mean changes and 95% confidence intervals. P values are for the adjusted time-by-subgroup interaction from linear mixed-effects models with fixed effects for time, subgroup, and their interaction and a patient-specific random intercept. Models were adjusted for age, sex, body mass index, changes in systolic blood pressure and heart rate, and intraprocedural crystalloid volume (mL/kg). Abbreviations: HR, heart rate; LAA, left atrial appendage; LAP, left atrial pressure; LASI, left atrial stiffness index; LAVI, left atrial volume index; PFA, pulsed field ablation; PVI, pulmonary vein isolation

**Figure 5.**
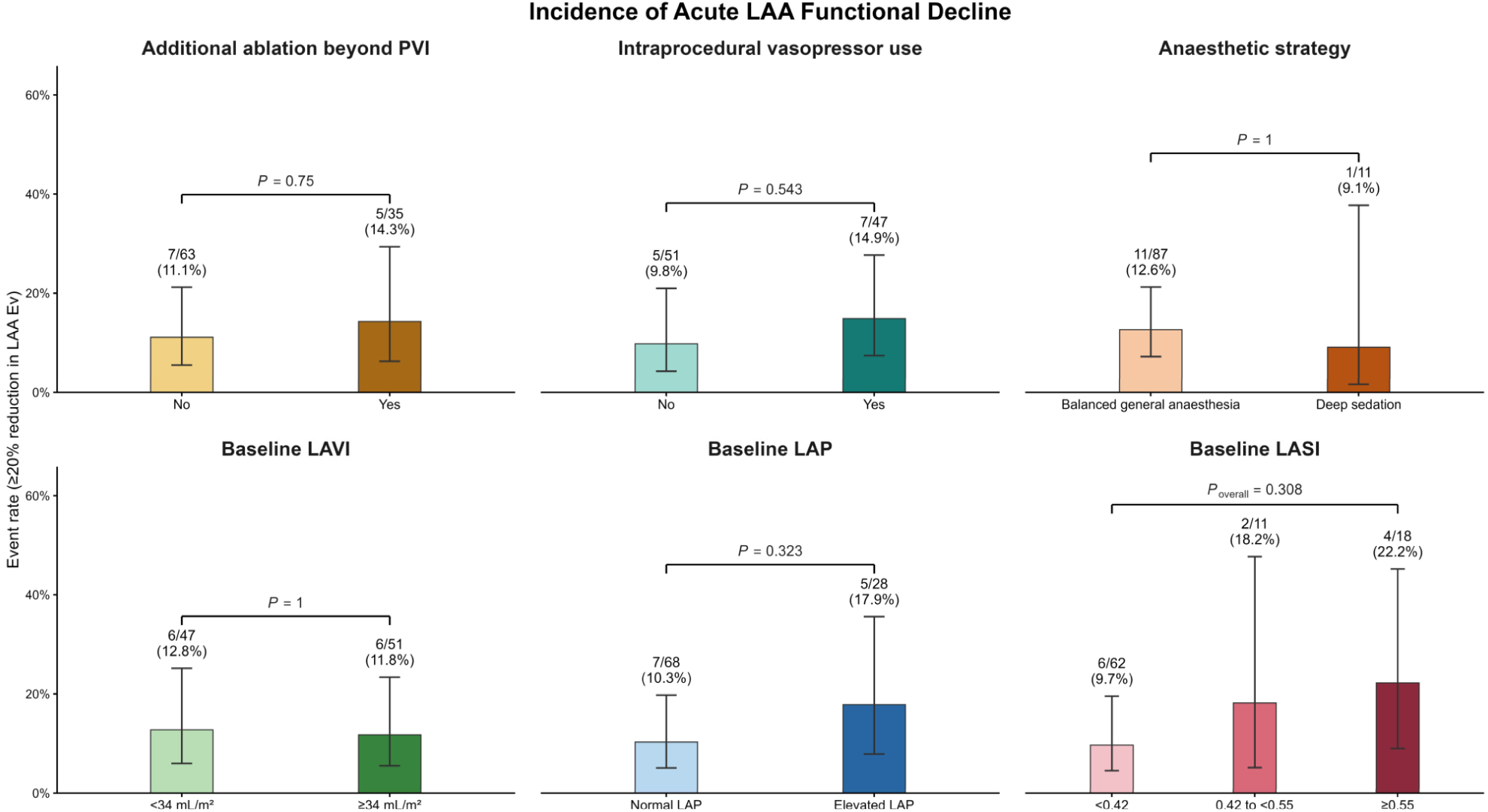
Incidence of acute LAA functional decline across procedural, anaesthetic, and echocardiographic subgroups. Acute LAA functional decline was defined as a ≥20% reduction in LAA emptying velocity from baseline. Bars show event proportions, labels report events/N (%), and error bars indicate Wilson 95% confidence intervals. Between-group P values were calculated using Fisher’s exact tests. Abbreviations: LAA, left atrial appendage; LAP, left atrial pressure; LASI, left atrial stiffness index; LAVI, left atrial volume index; PFA, pulsed field ablation; PVI, pulmonary vein isolation.

Among echocardiographic subgroups, mean changes in LAAEv were 6.3 and 4.6 cm/s in patients with baseline LAVI <34 and ≥34 mL/m², respectively (P for interaction = 0.700). Patients with elevated estimated LA pressure showed a numerically smaller increase than those with normal LA pressure (1.6 vs. 7.2 cm/s; P for interaction = 0.178). Across increasing LASI categories (<0.42, 0.42 to <0.55, and ≥0.55), mean changes were 6.2, −2.2, and 3.1 cm/s, respectively (P for interaction = 0.448). No statistically significant time-by-subgroup interactions were observed (Figure 4).

Marked acute reduction in LAAEv (≥20% from baseline) occurred in 12 of 98 patients (12.2%). The incidence did not differ significantly according to additional ablation beyond PVI (11.1% vs. 14.3%; P = 0.750), intraprocedural vasopressor use (9.8% vs. 14.9%; P = 0.543), or anaesthetic strategy (12.6% vs. 9.1%; P = 1.000). Similarly, no significant differences were observed across baseline LAVI (P = 1.000), estimated LA pressure (P = 0.323), or LASI categories (P = 0.308) (Figure 5).

## Discussion

In this prospective study using paired intraprocedural ICE, LAAEv and LAAFv both increased immediately after PFA, whereas transmitral A-wave velocity did not change significantly. Nevertheless, 12.2% of patients showed a marked acute reduction in LAAEv.

Several anatomical and procedural considerations suggest that PFA delivered near the left pulmonary veins could potentially affect LAA mechanical function. These include energy delivery along the Coumadin ridge^26^, and possible proximity of the pentaspline catheter to the LAA orifice during left-sided pulmonary vein ablation^27^. Our study, however, showed no overall acute impairment of LAA flow velocity, suggesting that LAA flow might not be adversely affected by PFA. Interestingly, LAA velocities increased immediately after PFA, which might be explained by several haemodynamic mechanisms.

LAA flow velocity is a dynamic, load-dependent physiological parameter rather than a direct measure of intrinsic appendage contractility^28, 29^. Experimental studies have shown that LAA filling and emptying are influenced not only by appendage contraction and relaxation but also by LA pressure and LV systolic and diastolic function^28, 29^. Consequently, acute changes in loading conditions and atrioventricular filling may alter LAA flow velocity even in the absence of a change in intrinsic LAA contractility^29^. This is particularly relevant intraprocedurally, when heart rate, blood pressure, and loading conditions may change over a short period.

Although heart rate and systolic blood pressure increased during the procedure, neither change was significantly associated with the acute change in LAA emptying velocity in the baseline-adjusted ANCOVA model. Intraprocedural vasopressor use was also not significantly associated with the observed change. Furthermore, exploratory subgroup analyses showed no significant differences in LAA emptying velocity changes according to vasopressor use or anaesthetic strategy. These findings suggest that the observed increase in LAA emptying velocity was not clearly attributable to the measured haemodynamic changes or differences in anaesthetic management. Nevertheless, potential effects of loading conditions, anaesthetic agents, and intraprocedural fluid administration cannot be excluded.

The baseline LAA emptying velocities observed in our study (median, 35.45 cm/s) were relatively low compared with values commonly reported in TEE-based studies of patients with paroxysmal AF^30–32^. Several factors may contribute to this difference, including the intraprocedural anaesthetic setting^33, 34^, and technical variations between ICE and TEE Doppler measurements^35^. In particular, differences in transducer position, insonation angle, and Doppler sample-volume placement may influence the recorded velocities. Therefore, absolute LAA emptying velocities obtained using intraprocedural ICE may not be directly interchangeable with those measured using TEE under different examination conditions. Importantly, the present study focused on paired measurements obtained using the same imaging modality within the same procedure, rather than comparisons with established TEE reference values.

The increase in LAA flow velocity occurred alongside increases in HR, blood pressure, and E-wave velocity, a shorter EDT, and no significant change in A-wave velocity. The increase in HR would be expected to shorten the diastolic filling period, while procedural volume loading may have increased preload. Together, these changes could have increased the early diastolic LA–LV pressure gradient, contributing to the higher E velocity and shorter EDT observed after PFA. One possible explanation is that enhanced early-diastolic LA-to-LV filling may facilitate LA unloading and thereby favour LAA emptying into the LA. This potential coupling between LA unloading and LAA emptying also raises the question of whether acute changes in LA pressure may contribute to the observed LAA flow response.

Previous pressure–volume loop studies have shown that radiofrequency and cryoballoon ablation are followed by an acute increase in LA pressure, with radiofrequency ablation additionally associated with increased LA stiffness^36^. Markedly elevated LA pressure has, in turn, been associated with lower LAA emptying velocities^28, 35, 37^. However, LAA emptying velocity increased in most patients in the present study. Although LA pressure was not directly measured during the procedure, this finding raises the possibility that PFA may produce a different acute atrial pressure–volume response from that reported after thermal ablation. A more limited rise in LA pressure could preserve more favourable conditions for LAA emptying, whereas a marked elevation in LA pressure may have the opposite effect^35^. This interpretation remains hypothetical and requires direct simultaneous measurement of LA pressure and LAA flow during PFA.

In the absence of direct intraprocedural pressure measurements, we therefore explored whether baseline non-invasive markers of atrial loading and stiffness were associated with the acute LAA flow response. Baseline LA pressure was estimated noninvasively from transthoracic echocardiographic parameters according to the 2025 ASE recommendations, an approach subsequently shown to agree well with invasive haemodynamic measurements^24, 38^. We also examined LASI, which integrates E/e′ with LA reservoir strain and reflects LA stiffness and compliance^23^, using age-specific upper reference limits^25, 39^. Patients with normal estimated LA pressure or lower LASI showed a numerically greater increase in LAAEv and fewer marked acute reductions than those with elevated LA pressure or higher LASI, respectively. This pattern would be consistent with the hypothesis that patients with already elevated LA pressure or a stiffer LA have less capacity to accommodate an acute rise in preload. Although neither subgroup analysis showed a statistically significant interaction, the concordant directional patterns suggest that baseline atrial loading conditions and stiffness may influence the acute LAA flow response to PFA. These findings remain exploratory, particularly given that only 12 of 98 patients experienced a marked acute reduction, and require confirmation in larger studies.

Our findings should also be considered alongside prior studies of LAA function after rhythm-control interventions. Acute reductions in LAAEv have been reported after electrical cardioversion^40, 41^ and radiofrequency ablation of atrial flutter^42^, while Donal et al. evaluated LAA flow immediately after radiofrequency PVI in patients with chronic AF using intraprocedural ICE^43^. In these studies, however, pre- and post-treatment measurements were obtained during different rhythms, making it difficult to distinguish procedural effects from those of rhythm restoration. Conversely, studies performed several months after thermal ablation have generally shown preserved or increased LAA flow^44, 45^, but these later changes may reflect sinus-rhythm maintenance and atrial reverse remodelling. To our knowledge, this is the first study to assess paired LAA flow velocities immediately before and after PFA without a concomitant change in rhythm.

## Data Availability

The datasets generated and analyzed during the current study are not publicly available due to patient privacy and institutional restrictions but may be made available from the corresponding author upon reasonable request, subject to institutional approval and applicable data protection regulations.

## Clinical Implications

Although the overall increase in LAA flow is reassuring, the marked acute reduction observed in 12.2% of patients should not be overlooked. Reduced LAA flow is associated with impaired appendage washout and thrombus formation, but the clinical significance of an acute reduction immediately after PFA remains unknown. Further studies are needed to determine whether these acute changes persist and whether they are associated with thromboembolic risk.

## Limitations

Several limitations should be acknowledged. First, this was a single-centre observational study without a thermal ablation control group; therefore, whether the observed changes are specific to PFA cannot be determined. Second, LA pressure was not measured invasively, and the proposed contribution of acute changes in loading conditions remains hypothetical. Baseline LA pressure was estimated using echocardiographic parameters and cannot substitute for direct intraprocedural pressure measurements. Third, LAA flow was assessed using ICE rather than TEE. Although paired measurements were obtained using a consistent imaging approach, Doppler velocities remain dependent on imaging plane and insonation angle. Fourth, measurements were obtained only immediately before and after PFA. We therefore cannot determine the duration of the observed changes or whether patients with marked acute reductions subsequently recovered. Fifth, the ≥20% reduction in LAAEv was used to characterize a marked acute decline and has not been validated as a threshold for adverse clinical outcomes. Moreover, only 12 patients met this definition, limiting statistical power for subgroup analyses. Sixth, measurements obtained at the end of the procedure may not capture delayed or maximal atrial stunning. Finally, the study was not designed to assess thrombus formation or thromboembolic events, and the clinical significance of these acute changes remains to be established.

## Conclusion

In patients undergoing PFA for AF, LAAEv and LAAFv increased immediately after ablation. These findings indicate that PFA does not produce a uniform acute impairment of LAA flow, although a minority of patients showed a marked acute reduction. The mechanisms and clinical significance of these acute changes require further investigation.

## Funding

This work received funding approval under the Junior and Pilot Research Projects (JPV) 2027 programme of University Hospital Královské Vinohrady, Prague, Czech Republic (project No. 13_JPV27).

## Disclosures

The authors have reported that they have no relationships relevant to the contents of this paper to disclose.

---

**Supplementary Table 1.**
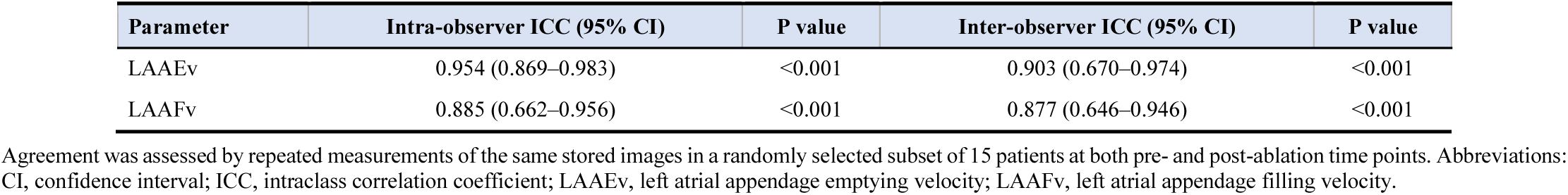
Observer agreement for LAA Doppler measurements. Agreement was assessed by repeated measurements of the same stored images in a randomly selected subset of 15 patients at both pre- and post-ablation time points. Abbreviations: CI, confidence interval; ICC, intraclass correlation coefficient; LAAEv, left atrial appendage emptying velocity; LAAFv, left atrial appendage filling velocity.

## Notes

### Competing Interest Statement

The authors have declared no competing interest.

### Author Declarations

The study was conducted in accordance with the Declaration of Helsinki and was approved by the Ethics Committee for Clinical Trials of University Hospital Královské Vinohrady (approval No. EK-VP/73/00/2025)

